# Multimodal AI-Based Risk Stratification for Distant Metastasis in Nasopharyngeal Carcinoma

**DOI:** 10.1101/2025.01.28.25321109

**Authors:** Jiayu Zhou, Made Satria Wibawa, Ruoyu Wang, Ying Deng, Haoyang Huang, Zhuoying Luo, Yue Xia, Xiang Guo, Lawrence S. Young, Kwok-Wai Lo, Nasir Rajpoot, Xing Lv

**Author notes:** joint corresponding authors: Nasir Rajpoot, Xiang Guo. joint first authors (Jiayu Zhou, Made Satria Wibawa).

## Abstract

**Background:** The TNM staging system is the primary tool for treatment decisions in nasopharyngeal carcinoma (NPC). However, therapeutic outcomes vary considerably between patients, and guidelines for the management of distant metastasis treatment remain limited. This study aimed to develop and validate a deep learning-based risk score to predict NPC survival.

**Methods:** We developed graph for nasopharyngeal carcinoma (GNPC) risk score, a multimodal deep learning based digital score incorporating signals from both Haematoxylin and Eosin (H&E)-stained tissue slides and clinical information. Digitised images of NPC tissue slides were represented as graphs to capture spatial context and tumour heterogeneity. The proposed GNPC score was developed and validated on 1,949 patients from two independent cohorts.

**Results:** The GNPC score successfully stratified patients in both cohorts, achieving statistically significant results for distant metastasis (p < 0.001), overall survival (p < 0.01) and local recurrence (p < 0.05). Further downstream analyses of morphological characteristics, molecular features, and genomic profiles identified several factors associated with GNPC score-based risk groups.

**Conclusion:** The proposed digital score demonstrates robust predictive performance for distant metastasis, overall survival, and local recurrence in NPC. These findings highlight its potential to assist with personalised treatment strategies and improve clinical management for NPC.

## Background

Nasopharyngeal carcinoma (NPC) is a malignant tumour of the head and neck that arises from the epithelium of the nasopharynx (1). Although it originates from similar tissue regions, NPC is distinctly different from other head and neck tumours. While relatively uncommon in North America and Europe, NPC is highly prevalent in Southeast Asia, where its incidence can reach up to 25 cases per 100,000 individuals (2). Globally, NPC is responsible for approximately 65,000 deaths each year (3).

NPC mortality has decreased with the widespread use of radiotherapy and chemotherapy, and the 5-year survival has generally improved (4,5). The primary challenges in treatment and leading causes of mortality are local recurrence and distant metastasis. Furthermore, additional 15% to 30% of those treated for locally advanced NPC will later develop local recurrence and/or distant metastasis (6,7).

The tumour-node-metastasis (TNM) staging system is the primary tool for prognostic prediction and risk stratification used to guide treatment decisions. However, outcomes for the same treatment can vary significantly among patients with the same TNM stage(8). The current first-line chemotherapy regimen for patients with metastatic NPC is a combination of gemcitabine and cisplatin (GP regimen). However, a subset of patients continues to experience poor prognoses despite this standard treatment (9,10). The variations in prognosis may be attributed to biological and tumour heterogeneity.

Therefore, it is crucial to develop new biomarkers that reflect tumour heterogeneity to better guide individualised treatment for patients. Due to the insufficiency of TNM staging system in guiding treatment for NPC, Epstein-Barr virus (EBV) DNA level has been proposed as a complementary biomarker. The development of NPC is strongly linked to Epstein–Barr virus (EBV) infection, though the precise mechanisms underlying its pathogenesis remain unclear (11). Elevated EBV DNA titres have been shown to originate from tumour cells and are widely used in clinical practice for diagnosing NPC, monitoring disease progression, and assessing post-treatment outcomes. However, a lack of standardisation in EBV DNA titre measurement across institutions has led to inconsistencies, making it challenging to compare results reliably (12).

Histological assessment remains the gold standard for cancer diagnosis and prognosis. Digital pathology has revolutionised this process by enabling histological tissues to be scanned into high-resolution images, known as whole slide images (WSIs). These images are rich in detail, capturing the morphological features of tumours, the complexity of the tumour microenvironment (TME), and tumour heterogeneity. Within WSIs, patterns in the TME — such as immune cell infiltration, stromal architecture, and spatial organisation —offer valuable insights into tumour biology.

WSIs have been widely utilised for the application of artificial intelligence (AI) in computational pathology (CPath) for various tasks, including detection of tumour structure (13,14), cancer grading (15,16), molecular pathway prediction (17), and cancer prognosis (18,19). Due to the large size of WSIs, slide-level tasks such as gene prediction and survival analysis often involve dividing the images into smaller patches for analysis using multiple instance learning (MIL) methods. While effective, MIL typically disregards spatial relationships between patches, which can result in the loss of critical contextual information.

Compared to other types of cancer, CPath studies in NPC that utilise WSIs as a data modality are far less common. In diagnostic tasks, WSIs have been used to differentiate between benign and malignant NPC tumours using deep learning (DL) approaches (20,21). Notably, in three-category classification tasks, the performance of DL models is comparable to that of senior pathologists (22) and surpasses the accuracy of junior and intermediate pathologists.

For prognostic tasks, microscopic pathological features extracted from WSIs have been employed to generate risk scores for progression-free survival in NPC patients (23).

Another study integrated clinical factors, histopathological features, and radiomic signatures to develop a multi-scale nomogram for predicting failure-free survival (24). Additionally, DL has been applied for the automated quantification of tumour-infiltrating lymphocytes (TILs) to stratify patient risk effectively (25). However, the generalisability of these studies is yet to be validated due to the small or single-centric cohorts.

This study aims to develop a novel biomarker for NPC by leveraging deep learning (DL) to integrate clinical information and histological images within a unified framework. We propose a graph neural network model for NPC (GNPC) risk score which takes graph representations of WSIs and combines features from a foundation model with morphological characteristics to represent the tumour microenvironment (TME). The graph comprises multiple disjoint subgraphs, each representing individual slides or distinct tissue regions, thereby effectively capturing tumour heterogeneity within each patient. Additionally, genomic data were utilised to further investigate the results of the GNPC score-based risk stratification, analysing the association of specific genetic mutations with immune cell infiltration and tumour nuclear morphology, thereby elucidating the molecular mechanism underlying NPC risk stratification. We employed two independent cohorts to train and evaluate our model. To the best of our knowledge, this is the first multi-centric study to apply DL with WSIs for patient prognostication in NPC.

## Methods

### Data

We retrospectively collected whole slide images (WSIs) and clinical data from two cohorts: the Sun Yat-sen University Cancer Centre (SYSUCC; n = 2,072 patients) and The Chinese University of Hong Kong (CUHK; n = 145 patients), with a total of 2,428 WSIs. All NPC tumours from CUHK cohort were collected by endoscopy or surgery at the Chinese University of Hong Kong, Hong Kong, SAR. Cases from SYSUCC cohort were diagnosed and treated between January 2011 and December 2018 at Sun Yat-sen University Cancer Center (Guangzhou, China). The median follow-up duration in the SYSUCC cohort was 76 months (range: 1–135 months), while in the CUHK cohort, it was 55 months (range: 1–222 months). In both cohorts, male patients accounted for more than 70% of the cases. Detailed clinicopathological information of both cohorts is available in the supplementary table S1.

To ensure the quality and integrity of the WSIs, a quantitative review was conducted on all WSIs using HistoQC (26). WSIs that exhibited poor quality, such as those with significant blurriness, a high prevalence of image artifacts, or those containing only minimal tissue areas, were identified and excluded from the analysis. Patients without available follow-up data, those with incomplete clinical information (i.e. sex, cancer stage, or treatment history), and patients with non-primary tumours were excluded.

Following this data cleaning process, the final cohort included 1,849 patients from SYSUCC and 100 patients from CUHK. Detailed information on the data exclusion process is available in the supplementary Figure S1.

We incorporated molecular data (EBV DNA titre and LMP1 status) and genomic data into our downstream analyses. EBV DNA data were available only in the SYSUCC cohort, whereas LMP1 status and genomic data were available in the CUHK cohort. Plasma EBV DNA levels were quantified using quantitative polymerase chain reaction targeting the BAMHI-W region of the EBV genome prior to treatment. Results were reported as the concentration of EBV genome copies per millilitre of plasma. Details regarding the collection of LMP1 status and genomic data can be found in these studies (27,28).

Patients from the SYSUCC cohort were divided into a discovery set (80%) and an internal validation set (20%). All patients in the CUHK cohort were used as the external validation set. The GNPC model was trained on the discovery set and evaluated on both the internal and external validation sets. Hyperparameters of the models were optimised using cross-validation within the discovery set.

### Graph for nasopharyngeal carcinoma risk stratification (GNPC)

Patients in our cohort may have multiple WSIs, with each WSI potentially containing more than one tissue area. For each WSI, we extracted image patches from tissue regions at a resolution of 0.5 microns per pixel (mpp). Leveraging the power of foundation models —pretrained deep learning architectures renowned for their generalisability and robustness across diverse tasks— we extracted deep features from these image patches, enabling a rich representation of the underlying pathology. Additionally, nuclei within each patch were detected and classified.

As shown in Figure 1, we constructed graphs from nodes that combine deep features and morphological features of tumour and inflammatory nuclei. Nodes were connected via weighted edges based on the node similarity, thereby forming one graph per tissue area. Graphs from different tissue areas or WSIs were combined into a single large, disjointed graph. This approach allows for a holistic representation of the entire patient sample. It captures variability across different tissue areas, offering better understanding of the spatial and morphological heterogeneity of tumours and its microenvironments.

**Figure 1.**
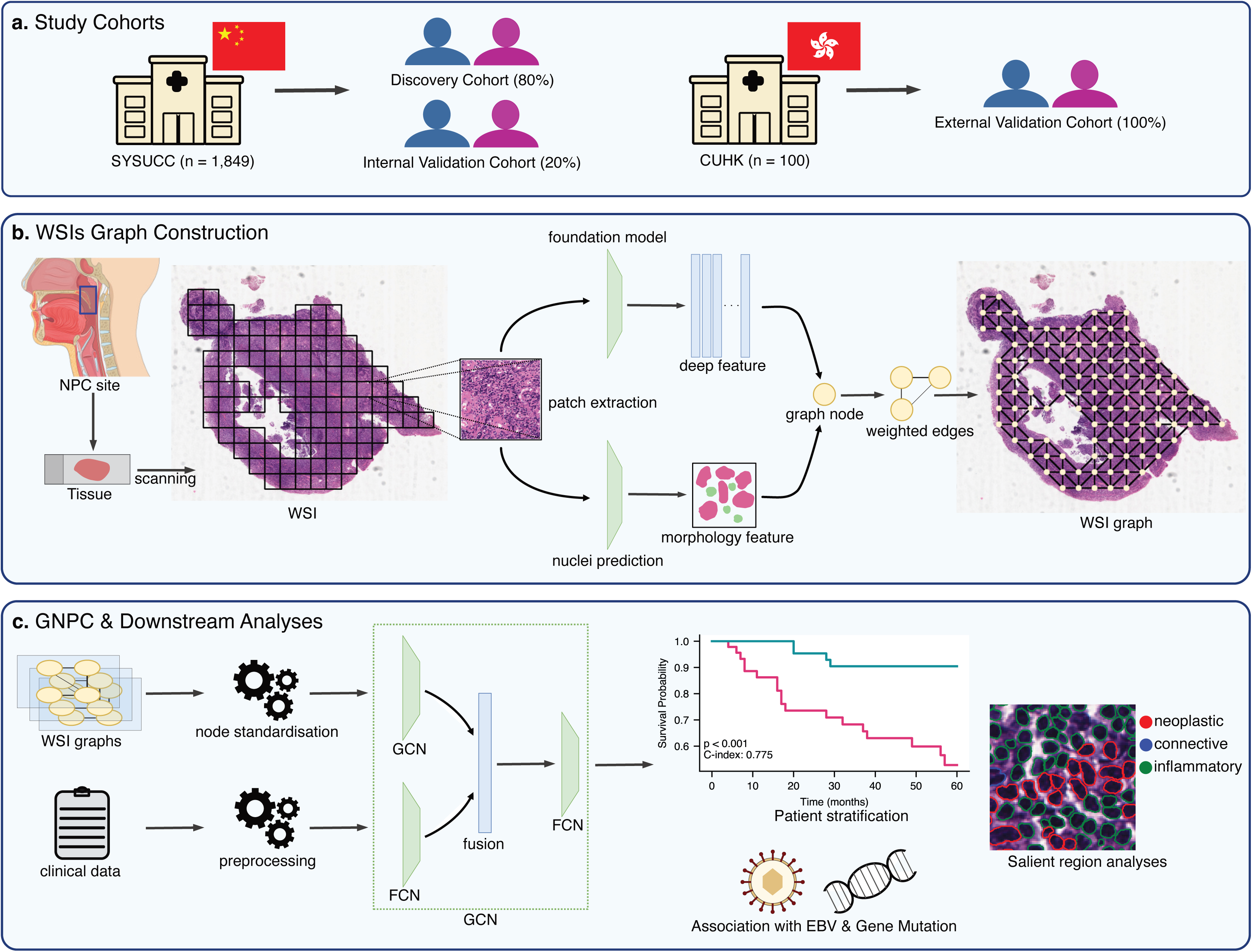
Main pipeline of the study. (a) Two cohorts were utilised as one discovery and two validations. (b) Graph were constructed from node of deep features and nuclei morphology features. (c) Multimodal model learns from WSI graphs and clinical data to generate risk score of NPC.

GNPC is a multimodal model wherein the latent embeddings of tissue patches and clinical data are amalgamated to generate a risk score. The latent embeddings from the graph convolutional network (GCN) and fully connected layer (FCN) were then concatenated and passed into the final layer. The final layer generated a continuous risk score, with a higher score corresponding to a higher risk of the event.

### Statistical analysis

The primary endpoint was distant metastasis-free survival (DMFS), calculated from the date of diagnosis to the date of the event. Additional endpoints included overall survival (OS) and local recurrence-free survival (LRFS). Survival data from both cohorts were right-censored, with a maximum observed follow-up period of 60 months (5 years).

Patients in the internal and external validation cohorts were stratified into low- and high-risk groups based on the median risk score derived from the discovery cohort. Patients with risk scores above the cutoff were classified as high-risk, while those with scores at or below the cutoff were designated as low-risk. These groups were compared using the log-rank test in Kaplan-Meier (KM) curve analysis. The concordance index (C-index) was employed to assess the predictive performance of the model.

Differences between groups were tested using Student’s t-test for continuous variables and Fisher’s exact test for categorical variables. Correlation analysis was performed using the Pearson correlation coefficient. Cox proportional hazards (Cox-PH) models were used to evaluate the prognostic value of the GNPC risk score and established risk factors in both univariate and multivariate analyses. A p-value < 0.05 was considered statistically significant, with significance levels denoted as p < 0.05 (*), p < 0.01 (**), and p < 0.001 (***). The main pipeline and downstream analyses were implemented in Python version 3.10. Statistical analyses were conducted using the lifelines package (https://lifelines.readthedocs.io/en/latest/) and the scikit-survival package (https://scikit-survival.readthedocs.io/en/stable/).

## Results

### Foundation models for survival prediction

We conducted comparative studies to assess the performance of the state-of-the-art foundation models (29–32) for survival prediction of NPC. Additionally, we used deep features from widely used deep learning model ResNet50 (33) as a baseline. All models were trained using 5-fold cross-validation derived from the discovery set. To evaluate model performance, we measured the C-index and the p-value from the log-rank test of the predicted risk scores. Risk scores in the validation set were stratified into low- and high-risk groups based on the median risk score value from the training set. We aggregated the results of 5-fold cross-validation by calculating the mean and standard deviation of the C-index and the median p-value from the log-rank test which are showed in the Figure. 2a.

**Figure 2.**
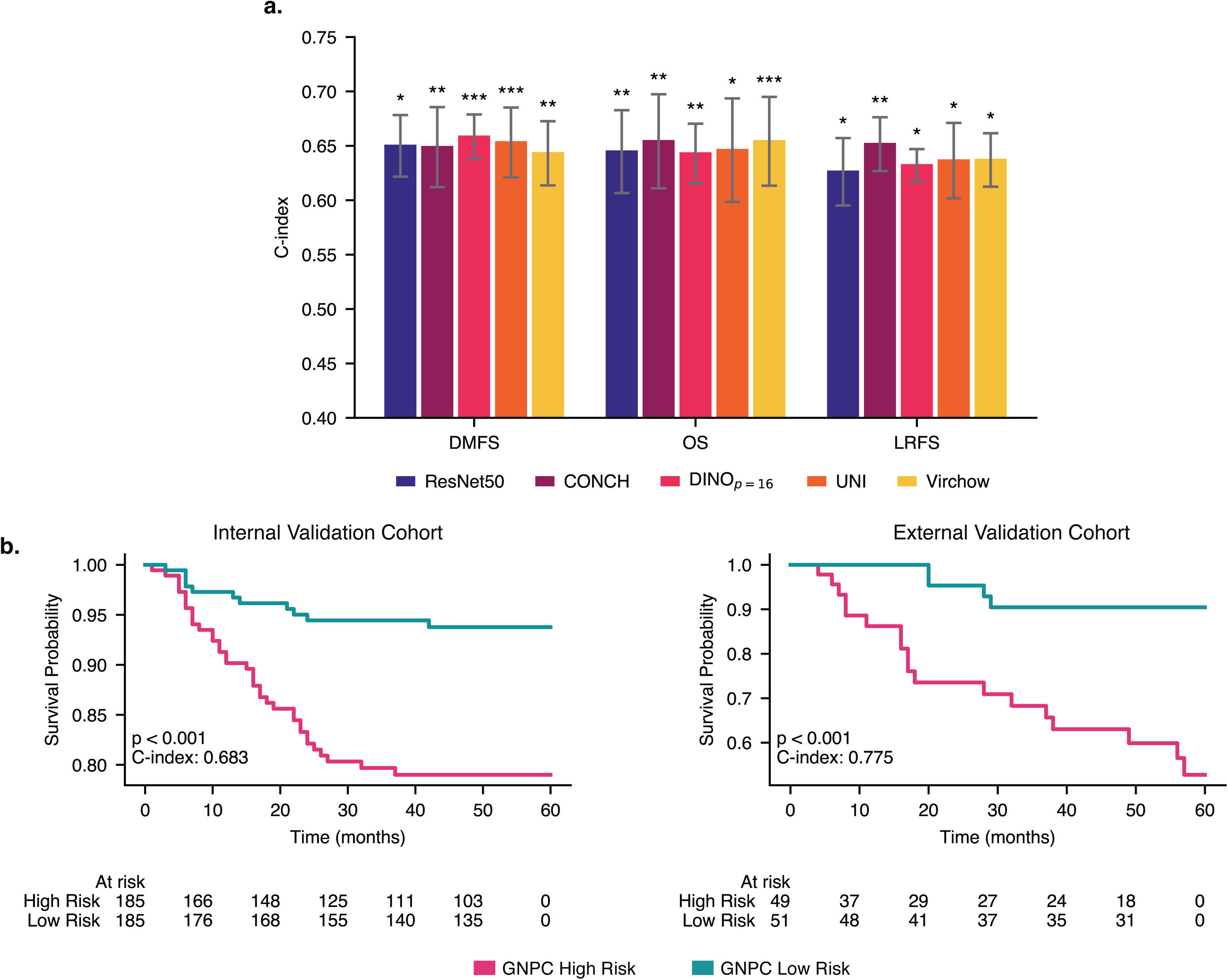
Survival prediction with GNPC. (a) Cross validation results for foundation models comparison. (b) GNPC evaluation with KM-curves on two validation cohorts for distant metastasis cases. Significance levels denoted as (*) (p < 0.05), ** (p < 0.01), and *** (p < 0.001).

Based on the median p-values across folds, all models in the experiments demonstrated statistically significant results. However, no model was particularly prominent, as all models show comparable predictive accuracy across the evaluated survival endpoints, with performance differences being relatively minor. In DMFS and OS, some foundation models deliver slightly worse performance compared to the baseline model, ResNet50, which was pretrained on non-histology images. To determine the deep features for the final model, we performed a top 3 voting based on the mean C-index across survival endpoints, resulting in the selection of CONCH (30) for deep feature extractor.

### Prognostic value of GNPC

Several patients in our cohorts had multiple H&E WSIs, and many of them contained multiple tissue regions. This intratumoural heterogeneity can act as a confounding factor in model prediction. GNPC captured all tissue regions and slides within each patient as one large graph consisted of disjointed subgraphs. Our model infers the risk score from this graph by focusing on the relevant regions based on attention pooling while preserving intratumoural heterogeneity.

The GNPC score achieved a C-index of 0.683 and 0.775 in the internal and external validation cohorts, respectively (Figure 2a). Additionally, a log-rank test yielded p < 0.001 for both cohorts, indicating that the GNPC score has statistically significant prognostic power in patient risk stratification. For 5-year OS, the GNPC score achieved C-indices of 0.657 and 0.814 in the internal and external validation cohorts, respectively (supplementary material Figure S4). The log-rank test indicated statistical significance, with p < 0.01 in the internal cohort and p < 0.001 in the external cohort (Figure 2b). Furthermore, for 5-year LRFS, our model achieved a C-index of 0.626 in the internal validation cohort (p < 0.01) and 0.647 in the external cohort (p < 0.05).

### Risk factors in NPC survival

Multiple factors, such as patient genetics, environmental influences, and viral infections, contribute to cancer progression in NPC. In this section, we examine the association between known NPC risk factors and the GNPC score with survival time across various survival endpoints.

Univariate CoxPH regression analysis showed that the GNPC score in both the internal validation cohort (p < 0.001) and the external validation cohort (p < 0.001) was significantly associated with poor prognosis in distant metastasis case (Figure 3). None of the other clinical factors were associated with distant metastasis prognosis in the internal validation cohort. Similar findings were observed for 5-year overall survival, where the GNPC score was significantly associated with poor prognosis in the internal validation cohort (p < 0.01) and the external validation cohort (p < 0.001). However, for cases of local recurrence, the GNPC score was significantly associated with poor prognosis only in the larger cohort (p < 0.01), with no significant association found in the external validation cohort (p = 0.125).

**Figure 3.**
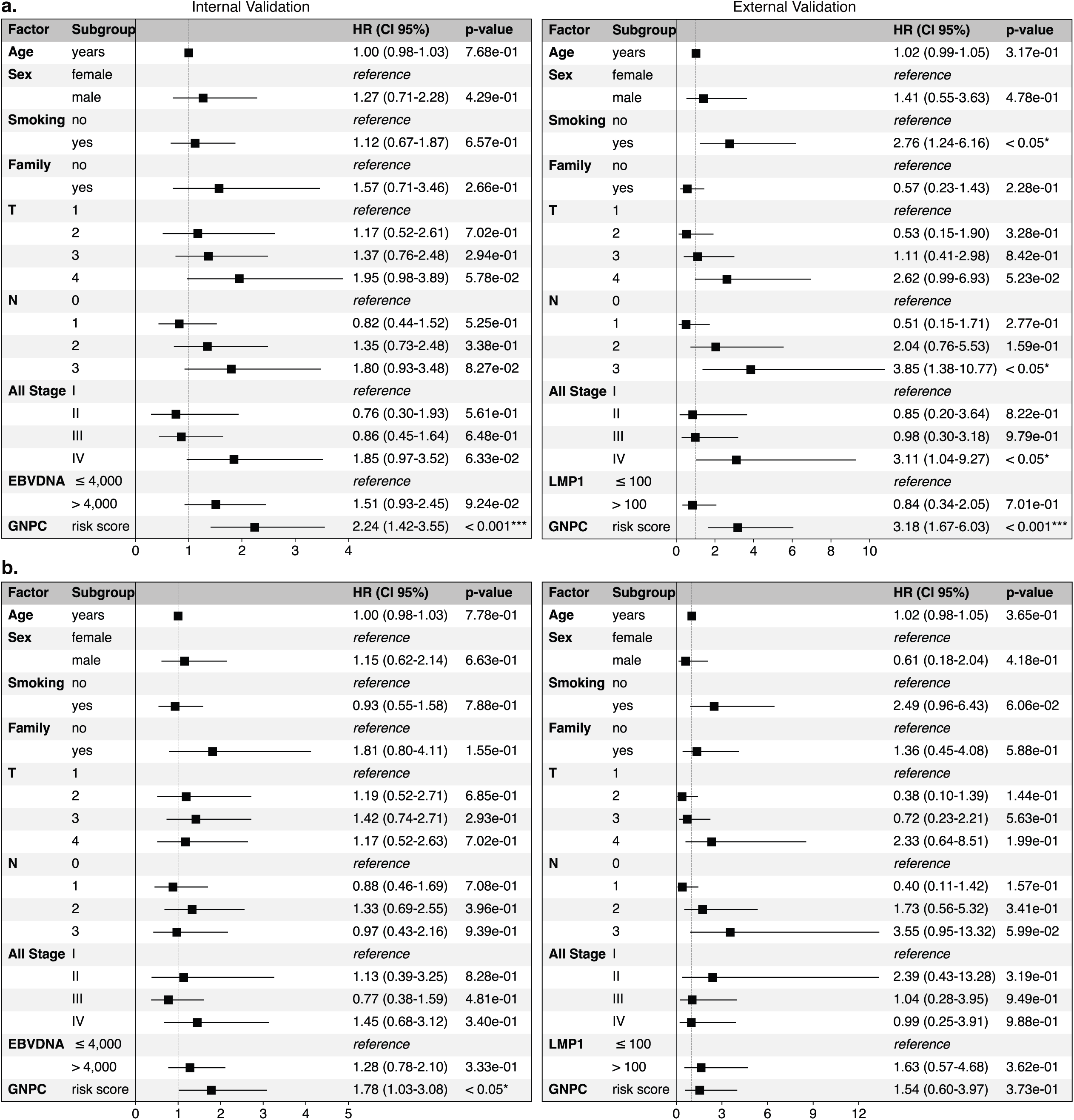
CoxPH analysis in distant metastasis for both validation cohort (a) Univariate and (b) multivariate. Significance levels denoted as (*) (p < 0.05), ** (p < 0.01), and *** (p < 0.001).

**Figure 4.**
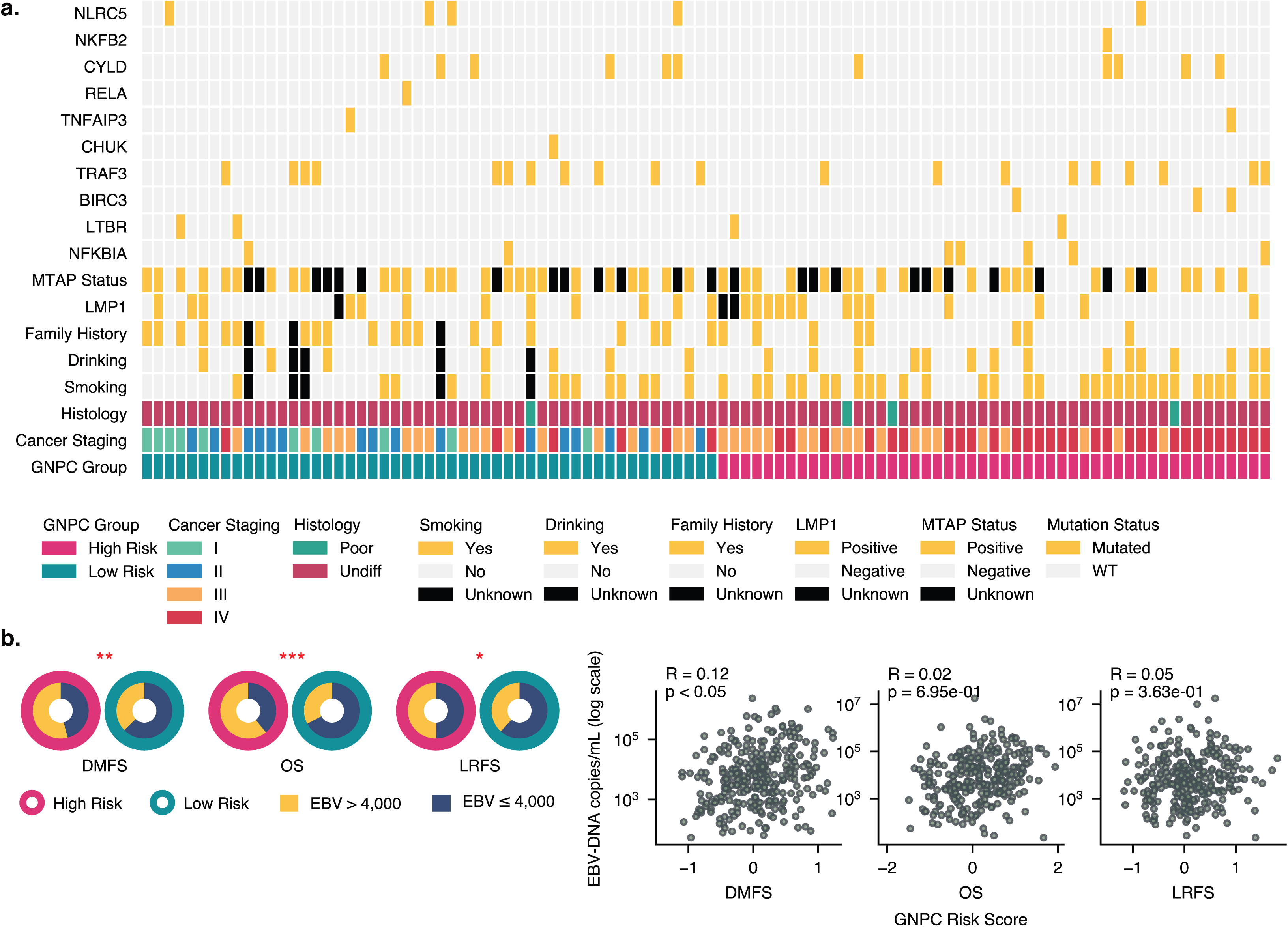
GNPC association with NPC risk factors in distant metastasis cases (a) Heatmap of risk factors and GNPC risk groups in the external validation cohort. (b) Distribution comparison (left) and correlation between EBV DNA copies/group and GNPC risk groups/scores in the internal validation cohort. Statistical significance denoted by red star. Significance levels denoted as (*) (p < 0.05), ** (p < 0.01), and *** (p < 0.001).

In multivariate CoxPH regression analysis, the GNPC score was found to be an independent risk factor in the internal validation cohort for DMFS (p < 0.001), OS (p < 0.05), and LRFS (p < 0.05), but not in the external validation cohort for DMFS (p = 0.373), OS (p = 0.197), or LRFS (p = 0.464). This discrepancy may be due to the limited sample size in the CUHK cohort.

### Association with risk factors and genomic alteration

Recent genomic studies indicate that NPC pathogenesis is characterised by frequent NF-𝜅B activation and immune evasion in 90% of cases. Investigating how specific gene alterations impact NPC prognosis could reveal new therapeutic opportunities for this disease.

We examined the distribution of genomic alterations within risk group of GNPC across survival endpoints, aiming to uncover molecular distinctions that may differentiate these groups. Although we found no statistically significant results, several genes showed consistent alteration patterns linked to risk group. For instance, alterations in NFKBIA and BIRC3 were more prevalent in the high-risk GNPC group, while NLRC5 alterations were more common in the low-risk group.

### GNPC risk score associated with EBV infection

Epstein-Barr virus (EBV) infection play a significant role in the development of NPC (1). Elevated levels of circulating cell-free EBV DNA were consistently found in the serum of NPC patients. Due to the strong correlation between EBV DNA levels and NPC progression, measurement of EBV DNA levels has become a tool for screening NPC (34,35).

We examined correlation between EBV DNA levels and the GNPC risk score in internal validation cohort. First, we stratified patients into low and high-EBV DNA groups using threshold of 4,000 (25,36) and compared the distribution of these groups within low and high-risk group of GNPC. Secondly, we measured correlation between GNPC score and EBV DNA levels. We consistently found a higher number of cases with elevated EBV DNA in the high-risk group across all survival endpoints, with statistically significant results (p<0.05). This suggests that patients in the high-risk GNPC category are more likely to have elevated EBV DNA levels, which may indicate a link between EBV viral load and increased risk as assessed by the GNPC model. This distribution pattern supports the hypothesis that EBV DNA could serve as an additional marker for identifying high-risk patients.

However, when directly correlating GNPC scores with EBV DNA levels, we observed only a weak but statistically significant relationship specifically in DMFS. This weak correlation (R = 0.12, p < 0.05) suggests that while there is a statistically meaningful association between EBV DNA and GNPC scores, the strength of this relationship is minimal. This implies that EBV DNA levels alone might not be a strong predictor of GNPC risk score across the entire cohort. Nevertheless, the statistical significance in DMFS highlights a possible subtle association between EBV DNA levels and the likelihood of distant metastasis, although the effect size is limited.

EBV infection in NPC encodes several viral oncogenes, including latent membrane protein 1 (LMP1). LMP1 exhibits immunomodulatory properties, influencing immune responses by altering the tumour microenvironment and promoting immune evasion. This protein is also involved in activating signalling pathways, such as NF-κB, that drive tumour growth and progression. Due to its pivotal role in NPC pathogenesis, LMP1 is recognised as a critical therapeutic target (34).

In this analysis, we investigate the association between GNPC scores and LMP1 expression in an external validation cohort respectively (supplementary material Figure S9). LMP1 expression was determined by immunohistochemical staining, assessed using a proportion score and an intensity score (37). Patients were stratified into absence/low and high LMP1 expression groups based on a H-score threshold of 100 (27), and we compared its distribution across GNPC risk groups. Although LMP1 is a known oncogene with important implication in NPC pathogenesis, we found no significant association between LMP1 expression levels and GNPC risk groups across all survival endpoints. This finding suggests that while LMP1 contributes to early NPC development, its expression does not appear to influence GNPC -based risk stratification outcomes.

### Salient region analyses

The pooling layer is a crucial component in GNNs, as it creates a refined representation of the graph by reducing and selecting the most relevant nodes for the prediction task. In our model, we utilised SAG pooling (38), which enables the model to focus on (or "attend to") specific nodes based on their relevance to the prediction task. This attention mechanism provides a way to interpret the model, allowing us to identify which parts of the tissues or slides are important for the prediction results and examine the tumour microenvironment (TME) in these salient regions. As shown in Figure 5a the top patches consisted of a high density of tumour cells, which is relevant to cancer progression.

**Figure 5.**
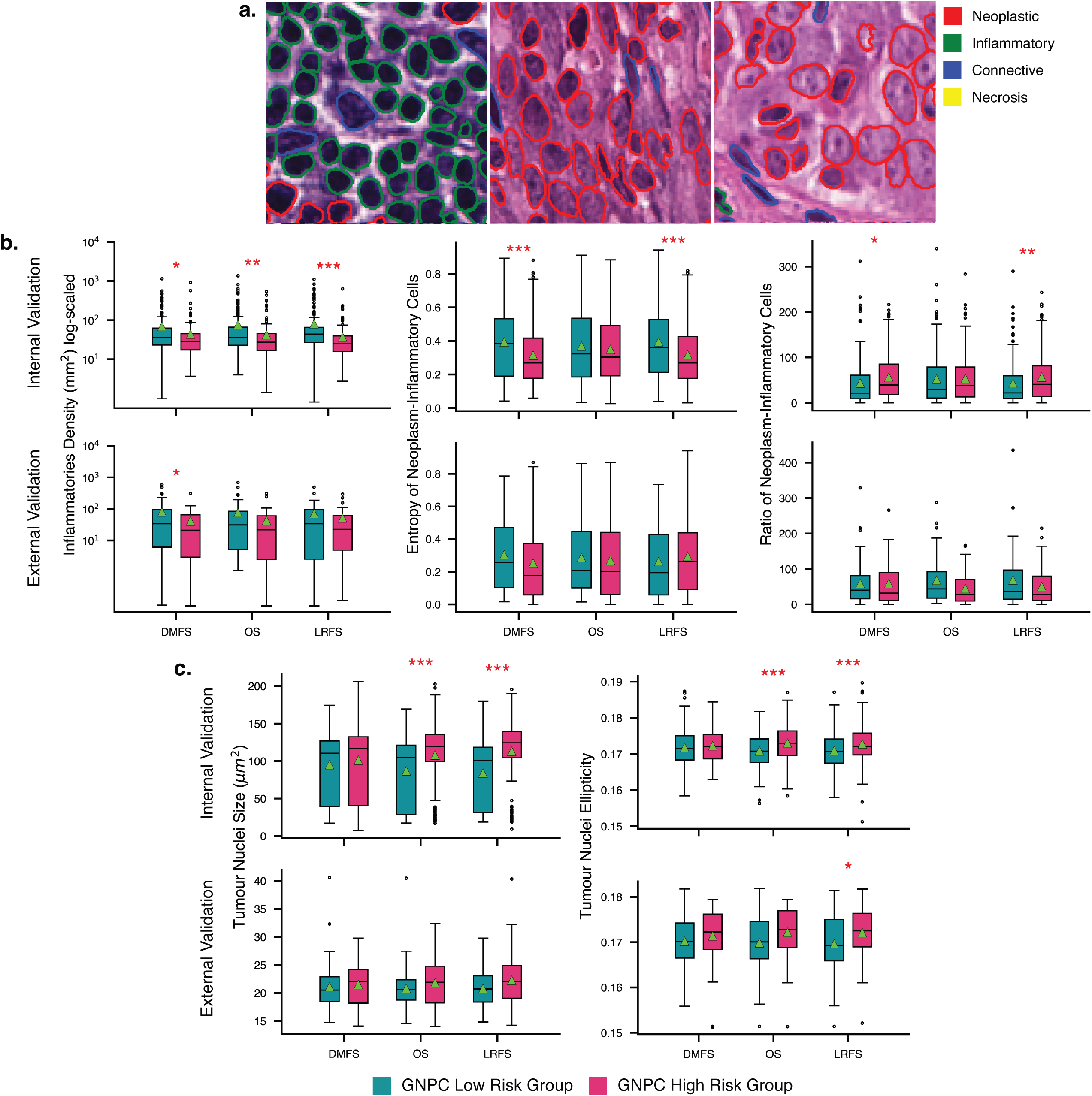
Salient regions in WSIs from top 10% patches. (a) Examples of nuclei composition in relevant image patches contributing to survival prediction. (b) Immune profile of GNPC risk groups across all survival endpoints. (c) Tumour nuclear pleomorphism across GNPC risk groups. Statistical significance is denoted by a red star. Significance levels denoted as (*) (p < 0.05), ** (p < 0.01), and *** (p < 0.001).

We selected the top 10% most relevant patches based on node’s attention score. Nuclei features in these relevant patches were then aggregated by calculating the mean at the slide level and subsequently at the patient level. Statistical differences between groups were assessed by comparing the mean of these aggregated results. From this salient region, we analysed the immune profile across GNPC risk groups by measuring the density of the inflammatory cells, as well as the entropy and ratio of tumour (neoplasm) cells to inflammatory cells. Density was defined as the abundance of inflammatory cells per mm^2^, while entropy and ratio described the heterogeneity of tumour and inflammatory cells. Together, these metrics reflect the immune activity. Additionally, we investigated tumour morphology, focusing on shape ellipticity. Ellipticity was quantified using eccentricity, where a higher value indicates greater deviation from a perfect circular shape.

### Higher density of immune cells in the lower risk group

NPC is characterised by a high presence of tumour-infiltrating lymphocytes (TILs), which have been associated with improved outcomes, including distant metastasis-free survival and overall survival (39). TILs play a crucial role in the onset, progression, invasion, and metastasis of NPC and have been suggested as a potential prognostic biomarker for the disease (40). We analysed inflammatory cells density in the top 10% of tumour regions, comparing low- and high-risk groups (Figure 5b). Our findings indicate that lower-risk groups tend to exhibit a higher average density of inflammatory cells infiltration across all survival endpoints in the validation cohort. This association was statistically significant across all survival endpoints in the internal validation cohort. However, in the external validation cohort, which had fewer cases, statistical significance was observed only for distant metastasis. Our model effectively captured the complex interplay between TILs and NPC progression, aligning with findings from previous studies (41–43).

### Increased nuclear pleomorphism in the high-risk group

It has been suggested that neoplastic spindle cells, prominently located at the invasive tumour front and within the surrounding stroma, are associated with poor prognosis in NPC (44). High proportion of spindle tumour cells in NPC indicates poor prognosis, including overall survival, clinical stage, lymphatic invasion, and recurrence, compared to the non-spindle subtypes (45,46). Using the same pipeline as our analysis on inflammatory cells infiltration, we measured tumour ellipticity at the patient level (Figure 5c). Based on our findings, high risk patient in both validation cohorts tend to have larger tumour nuclei in average. For the OS and LRFS case in the internal cohort, this pattern was statistically significant (p<0.05).

Nuclear ellipticity in tumour cells is indicated by an increased spindle-like shape of nuclei. Genetic instability in poor-prognosis tumours leads to abnormal cell division, which, among other effects, results in the formation of spindle cells. By examining tumour morphology in salient regions, we observed this phenomenon as well. On average, tumour morphology in the higher-risk group in both cohorts exhibit a more spindle shape compared to the lower-risk group. Additionally, this pattern is statistically significant in the OS case of the internal cohort and in the LRFS case across both cohorts.

### Association between immune characteristics and gene mutation in NPC

Lymphocyte infiltration has emerged as a critical factor influencing cancer progression and prognosis. Recent studies highlight that specific gene mutations can significantly affect immune cell dynamics within tumours (28,47), potentially altering lymphocytes count. To further investigate this relationship, we analysed lymphocyte/ inflammatory cells density across tumour regions with different gene mutation profiles.

We compared how lymphocyte density differs between groups of patients with non-mutated and mutated genes (Figure 6). Inflammatory density was measured by calculating the number of inflammatory cells nuclei in salient tumour regions. This analysis was conducted on an external validation (CUHK) cohort. Across all survival endpoints, we observed no significant difference in inflammatory cells density between groups, except for lymphotoxin-beta receptor (LTBR). Patients with an LTBR amplification exhibited a higher inflammatory cells density, with statistically significant results. LTBR is crucial in both development and regulation of immune system (48).Previous studies have demonstrated LTBR as a driver of nonconical NF-κB activation in NPC (28,49). The association between LTBR amplification and increased inflammatory cells suggests a potential link between LTBR-mediated nonconical NF-κB activation and immune cells recruitment within the TMEs.

**Figure 6.**
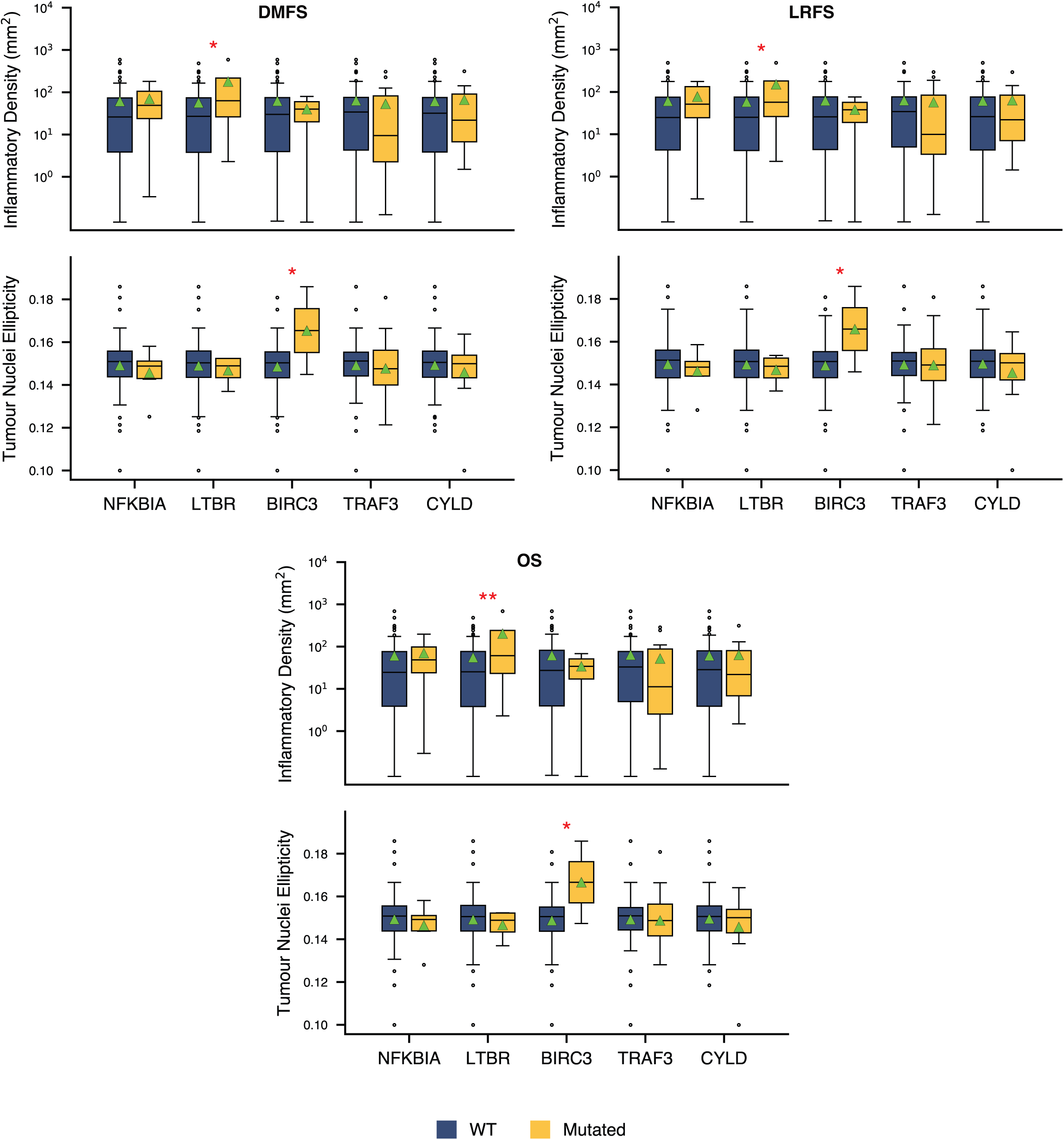
Association between gene alteration and immune cell density and tumour nuclei ellipticity across all survival endpoints. Statistical significance is denoted by a red star. Significance levels denoted as (*) (p < 0.05), ** (p < 0.01), and *** (p < 0.001).

### Specific gene mutation associated with spindle tumour cells

Gene mutations may drive cancer progression, often leading to observable changes in tumour cell morphology that may reflect underlying biological processes. Specifically, NF-𝜅B pathway which represents transcription factor involved in the regularisation of tumour proliferation and apoptosis (47). In this section, we examine how specific mutations correlate with variations in tumour cell structure, focusing on nuclear size and ellipticity.

By comparing morphologic traits across mutation profiles, our analysis reveals a distinctive pattern in tumour nuclei shape with genetic alterations of BIRC3. The proportion of tumour nuclei exhibiting a spindle shape is significantly higher (p < 0.05) in the group of patients with altered BIRC3 in the external validation cohort across all survival endpoints. However, there is no significant difference of between tumour cells size in various genetic alteration.

## Discussion

In this multicohort retrospective study, we introduce the digital GNPC risk score which based on multimodal graph learning for predicting distant metastasis risk in nasopharyngeal carcinoma (NPC). The GNPC risk score encodes complex interactions within the tumour microenvironment across slides and tissue areas into a unified graph by integrating foundation model features, cellular morphology, spatial location, and clinical information. Our model was trained and evaluated on a large cohort with a total of 1,949 patients. The score demonstrates strong performance and generalisability in both validation cohorts for predicting distant metastasis in NPC. Additionally, it also achieved significant results in risk stratification for overall survival and local recurrence. The consistent statistical significance observed in the log-rank tests across different survival endpoints highlights the score’s strong predictive capability for risk stratification. Notably, the higher C-index values in the external validation cohort suggest that the GNPC risk score is generalisable and effective beyond the original discovery set. These findings support the potential clinical utility of the score in accurately identifying high-risk patients who may benefit from closer monitoring or more aggressive therapeutic strategies, thereby providing valuable insights for individualized risk assessment and treatment planning.

Nearly 95% of individuals across the globe are asymptomatically infected with EBV (50,51). EBV infection plays a critical role in the pathogenesis of NPC, with multiple extrachromosomal copies of the EBV genome present in every tumour cell (52,53). Elevated EBV serological markers in NPC patients are associated with poor prognosis, making it a common practice to use EBV DNA as a prognostic marker to guide personalized treatment strategies. Using EBV DNA data from the internal validation cohort, we examined its association with the proposed GNPC risk score. We found patients with high EBV DNA levels (> 4,000 copies/mL) were associated with the high-risk group as per our risk score across all survival endpoints. Specifically, in distant metastasis cases, the GNPC risk score revealed a statistically significant association with EBV DNA levels.

Persistent EBV infection contributes to the progression of NPC by expressing viral proteins such as EBNA1 and the LMP proteins, which drive the oncogenic process as well as disrupting immune surveillance mechanisms (54,55). NPC is characterised by a significant tumour microenvironment consisting of stromal cells as a significant infiltration of immune cells. A higher infiltration of immune cells within the tumour area is associated with a better prognosis in NPC (56). In our analysis of salient regions of NPC, we discovered that the low-risk group demonstrates higher level of immune cell activity compared to the higher risk group of GNPC. This finding underscores the potential of GNPC as a biomarker for identifying immune-active NPC in EBV-infected cases, which may benefit from immune-targeted therapies. Despite, the important role of LMP1 in immune regulation of NPC, we did not observed association between LMP1 status with GNPC’s risk group. This may be due to the limited number of cases in the cohort with LMP1 data and other somatic gene alterations contributed to immune regulation and inflammation in the tumours. The correlation of LMP1 and genetic alterations with the GNPC requires further investigation in a large cohort of NPC samples in future.

Despite the widespread use of the WHO subtype system for clinical classification of NPC, a growing number of studies suggest that the current WHO classification is inadequate for predicting chemotherapy and radiotherapy outcomes (44,45,57).

Morphological characteristics of neoplastic cells have been proposed as a potential factor for improving NPC prognosis, with a higher proportion of neoplastic spindle cells linked to worse outcomes. By incorporating neoplastic nuclear morphology as node features in the graph, our model learns subtle morphological patterns in relation to survival outcomes. Our analysis of salient regions revealed that tumour nuclei in the high-risk group of GNPC have larger nuclei and exhibit a spindle shape compared to those in the low-risk group. These changes are believed to reflect increased cell plasticity associated with epithelial-to-mesenchymal transition (EMT) and epigenetic alteration during cancer progression. This indicates that while our model incorporates the WHO classification in its multimodal branch, it also aligns with recent studies highlighting the prognostic value of morphology of neoplastic spindle cells.

The development of NPC is influenced by a combination of host genetics, environmental factors, and EBV infection. Several key genetic and epigenetic events have been identified as contributors to tumour progression. Although no significant differences were observed in the GNPC risk group between mutated and non-mutated genes, certain genes, such as NFKBIA and BIRC3, were more frequently altered in the high-risk GNPC group across all survival endpoints. Our TME profiling in salient regions revealed a significantly higher immune cell density in altered LTBR group. Furthermore, tumour in the altered BIRC3 group exhibited more spindle-shaped cells.

These findings align with the role of LTBR activation and inactivation of BIRC2, BIRC3 and TRAF3 in the activation of noncanonical NF-kB pathway. Mutations in NFKBIA (IκBα) and BIRC3 (cIAP2) are believed to exert a more potent effect on NF-κB signalling and inflammation compared to other negative regulators of the pathway, such as TRAF3 and CYLD. This dysregulation may promote tumour survival and progression. In particular, the loss of BIRC3/cIAP2 function in NPC is hypothesised to drive activation of the noncanonical NF-κB pathway (40,47). Additionally, independent studies have identified loss-of-function mutations in NF-κB negative regulators, including NFKBIA, in NPC tumours (58). Collectively, these findings suggest that dysregulated NF-κB signalling, driven by genetic alterations, is a central mechanism in NPC pathogenesis.

Our study offers several key contributions. To the best of our knowledge, this is one of the largest DL-based prognosis studies in NPC that utilises histological images to date. Our multimodal approach leverages prognostic information from various factors. The graph-based methodology enables us to capture context from spatial relationships in the TME while preserving intratumoural heterogeneity. Furthermore, our model identifies patterns in biological mechanisms, including nuclear pleomorphism, EBV infection, and their correlations with immune response.

One limitation of our study is that molecular data, such as EBV DNA, LMP1, and genomic alterations, was available only for a single cohort rather than across both cohorts. This limitation restricted our ability to generalise findings related to the molecular profiles of NPC. Additionally, the data in this study was acquired solely from cohorts in Asia, where NPC is highly prevalent. This geographic concentration may limit the generalisability of our findings to other populations, such as those in Europe or the US, where NPC characteristics and risk factors may differ. Further validation with diverse cohorts would be beneficial to confirm the applicability of our model across various demographic and regional backgrounds.

## Data Availability

All data produced in the present study are available upon reasonable request to the authors

## Acknowledgement

This study is fully supported by a PhD scholarship to MSW funded by Indonesia Endowment Fund for Education (LPDP), Ministry of Finance, Republic of Indonesia under grant number Ref: S-575/LPDP.4/2020.

## Ethical Statement

Informed consent was obtained from all CUHK participants in accordance with institutional clinical research approvals. Data from SYSUCC and all procedures were conducted in accordance with ethical principles (B2023-381-01).

## Author Contributions

MSW, JZ, RW, KWL, LSY, NMR designed the study with the help of all co-authors. MSW, NMR developed the computational methods. MSW wrote the code and carried out the experiments. JW, KWL, YD, HH, ZL, YX, XG, XL obtained the ethical approval and retrieved the histological and clinical data from Sun Yat-sen University Cancer Center and The Chinese University of Hong-Kong. MSW prepared the original draft. All authors have read and agreed to this version of the manuscript.

## Code Availability

Source code is made publicly available upon the publication, subject to intellectual property constrains (https://github.com/mdsatria/GNPC).

## Data Availability

Data can be made available upon request to the corresponding authors.

## Funding

MSW is funded by PhD scholarship, Indonesia Endowment Fund for Education (LPDP), Ministry of Finance, Republic of Indonesia under grant number Ref: S-575/LPDP.4/2020.

## Competing Interest

NMR is the co-founder, CEO and CSO of Histofy Ltd., UK. He is also the GSK Chair of Computational Pathology and is in receipt of research funding from GSK and AstraZeneca.

## Notes

### Author Declarations

Informed consent was obtained from all CUHK participants in accordance with institutional clinical research approvals. Data from SYSUCC and all procedures were conducted in accordance with ethical principles (B2023-381-01). CUHK cohort: The Joint Chinese University of Hong Kong - New Territories East Cluster Clinical Research Ethics Committee approved this study. Ethical approval was granted. SYSUCC cohort: The Research Ethics Committee of the Sun Yat-sen University Cancer Center approved this study. Ethical approval was granted.

