## supplementary material for "Multimodal AI-Based Risk Stratification for Distant Metastasis in Nasopharyngeal Carcinoma"

#### GNPC Pipeline

##### *Image patches as graph nodes*

Separation between tissue and non-tissue areas was performed at a lower magnification using Otsu thresholding for image segmentation. Following image segmentation, small objects and holes were removed using scikit-image to eliminate image artifacts and improve quality.

Non-overlapping patches, each sized 256x256 pixels at 20x magnification or 0.5 mpp, were extracted from all tissue regions in the WSIs. These patches are the representation of nodes in the WSI graph. Node features, consisting of deep features and nuclei morphology were extracted in the subsequent data pipeline.

##### *Deep feature extraction*

Pathology images are inherently distinct from natural images, as they tend to exhibit low colour variation and lack canonical orientation. These characteristics can reduce the effectiveness of pretrained features, originally trained on natural images, when applied in the pathology domain. The advancement of self-supervised learning (SSL) enables the utilization of unlabelled data, which is particularly valuable in pathology, where annotations are costly to obtain. SSL can leverage unannotated pathology images to generate richer and more meaningful feature representations.

Owing to this, we employed four state-of-the-art SSL models, trained on histopathology data, to extract deep features from the patches. The SSL models used were VIRCHOW (1), CONCH (2), UNI (3) and DINO (4). For comparison, we also used deep features extracted from the ResNet50 model, pretrained on ImageNet. The feature vector lengths for each model were: 512 for CONCH, 384 for DINO, 1024 for UNI, 2048 for ResNet50, and 2560 for VIRCHOW. None of the data from this study were used to train the models for deep feature extraction, ensuring no data leakage occurred.

##### *Nuclei morphology features*

NPC is characterised by significant infiltration of leukocytes into the tumour microenvironment (TME). The abundance and composition of tumour-infiltrating lymphocytes (TILs) have been shown to correlate with patient prognosis in NPC(5). Additionally, the morphological characteristics of tumour cells have been reported to contribute to the aggressiveness of NPC(6) . For these reasons, we included morphological features representing the size and shape of both tumour and lymphocyte nuclei in the image patches as node features.

Size of nuclei was quantified using measurements such as contour area, convex-hull area, equivalent diameter, minor axis length, major axis length, perimeter, bounding box area, and radius. Furthermore, nuclear shape was quantified using features such as roundness, eccentricity, solidity, and orientation. In total, 12 morphological features were analysed.

These features were extracted from both lymphocyte and tumour nuclei in each image patch. To aggregate the features at the patch level, we calculated the mean, standard deviation, kurtosis, range, and skewness for each nuclear type. This resulted in a morphological feature vector with a total length of 144 (12 features × 2 nuclei types × 6 statistical measures).

#### *Clinical features*

To incorporate clinical information into our multimodal GNN prognostic model, we used 7 clinical features: age, sex, smoking status, family history of cancer, T stage, N stage, and overall stage. We applied one-hot encoding for categorical features and standardisation for numerical features before passing them to the model.

$$Z = \frac{X - \mu}{\sigma}$$

#### *Graph construction*

A graph is a data structure consisting of nodes (vertices) and edges (connections). This structure is well-suited for modelling large images, such as whole slide images (WSIs). The WSI graph was constructed by connecting nodes, each represented by image patches. The node features consisted of vectors obtained from the concatenation of deep features and morphological features. Connections between nodes were established using Delaunay triangulation, with a threshold of 2,000 pixels. This threshold was empirically defined to limit connections between nodes within a single tissue area. Furthermore, to capture spatial relationships and heterogeneity in the tumour microenvironment, edges were weighted based on the cosine similarity of node features. Cosine similarity assigns a score between 0 and 1 to the feature vectors, where 1 indicates that the vectors are identical, and 0 indicates they are completely unrelated. The edge weights capture both spatial relationships and heterogeneity in the tumour microenvironment. Several patients in the SYSUCC cohort had multiple slides, and some slides contained multiple tissue areas. To accommodate this complexity, we combined all tissue areas for each patient into one large graph, consisting of disjointed smaller graphs.

#### *GNN model*

In a graph of $G$, the graph is represented by $G = (V, E)$, where $V$ denotes the set of nodes and $E$ represents the set of edges that link these nodes. The graph structure is encoded in adjacency matrix $A$, where $A_{ij}$ is equal to 1 if node $i$ and $j$ are connected, and $A_{ij}$ is equal to 0 if otherwise. In our WSI graph $G$, the $V$ was image patches which represented by concatenation of deep features and morphological features of tumour and lymphocytes nuclei. $E$ was constructed using Delaunay triangulation and weighted by a cosine similarity function. This approach captures tissue structure, which is important for representing the tumour microenvironment. In addition, the weighted $E$ enable the model to highlight patterns and relationship in regions while reduce impact of irrelevant regions.

A GNN learns from graph by updating each node's feature representation through the aggregation of information from its neighbour, a process formally known as message passing. Various message passing algorithm have been proposed. In this study, we used Graph Convolution Networks (GCN)(7) proposed by Kipf and Welling for message passing.

In GCN, node representations are updated through a convolution operation defined as follow:

$$H^{(l+1)} = \sigma\left(\tilde{A}H^{(l)}W^{(l)}\right)$$

where $H^{(l)}$ is the feature matrix in the $l^{th}$ layer and $H^{(0)} = X$. In this study, $\sigma(.)$ denotes an activation function, specifically Leaky-ReLU(8), which is more suitable than standard ReLU for complex and diverse structure such as graphs. $W^{(l)}$ is a learnable weight matrix for layer $l$.

Each successive GCN layers learn new feature matrix $H^{(l)}$ that represents each node along with its neighbourhood information before aggregation operation in pooling layer. We used self-attention graph (SAGPooling)(9) with hierarchical architecture as pooling layer to aggregate node information. SAGPooling computes an attention score $s$ for each node $V$ using linear transformation followed by non-linear activation, specifically $tanh$ in this study.

$$s = tanh\left(H^{(l)} \cdot w\right)$$

The GNN in this study comprised of four layers, each of which includes a GCN and SAGPooling layer. The output of each block is aggregated in the readout step using the operation

$$h^{(j)} = \frac{1}{N}\sum_{i=1}^{N} x_i \,||max_{i=i}^{N} x_i$$

and is summarised by summation operation in final readout layer. This latent representation of graph is then concatenated with latent representation of clinical information. The combined embedding is then passed to the final fully connected layer, which has a single output node.

##### *Loss function*

The model was trained using regularised negative log-likelihood (NLL) loss derived from the partial likelihood of the Cox proportional hazard model. The Cox model assumes a hazard function at time $t$ for individual with covariates $x$, denoted as $h(t|x)$, is proportional to a baseline hazard function $h_0(t)$.

Let $l(\theta)$ denote the NLL loss:

$$l(\theta) := -\frac{1}{N_{E=1}} \sum_{i:E_i=1} \left( \widehat{h_\theta}(x_i) - \log \sum_{j \in R(T_i)} e^{\widehat{h_\theta}(x_j)} \right) + \lambda \cdot |\theta|_2^2$$

where $N_E$ is the number of observed (uncensored) events $(E = 1)$. Here, $\widehat{h_\theta}(x_i)$ is the predicted risk score of individuals $i$ under the model parameters $\theta$, as represented by features vector $x_i$. The risk set $R(T_i)$ consists of all individuals $j$ who are still at risk at time $T_i$, including those whose event or censoring time satisfies $T_j \geq T_i$.

The objective is to minimise the negative log-partial likelihood under the assumption that higher risk scores correspond to worse outcomes (i.e., shorter time to the event). For each observed event $(E_i = 1)$, the risk score $\widehat{h_\theta}(x_i)$ is compared to the normalised cumulative risk of individuals in the risk set $R(T_i)$. Finally, the loss is regularised using L2 regularisation with strength $\lambda$, which penalises large parameters to reduce overfitting and improve generalisation.

##### *Multimodal GNN for NPC survival (GNPC)*

GNPC is a multimodal model comprising two concurrent networks: a graph neural network (GNN) and a fully connected network (FCN). The latent embeddings from each network were concatenated into an intermediate network before being passed into the final fully connected network.

For the GNN, we used a graph convolutional network (GCN) as the convolutional layer, followed by a Leaky-ReLU non-linear activation layer. All FCN layers utilised the scaled exponential linear unit (SELU) as the activation function. To mitigate overfitting, dropout with a rate of 0.4 was applied to each layer of the FCN. Additionally, 1D batch normalisation was employed in the FCN to stabilise training.

**Figure S1 GNPC architecture**

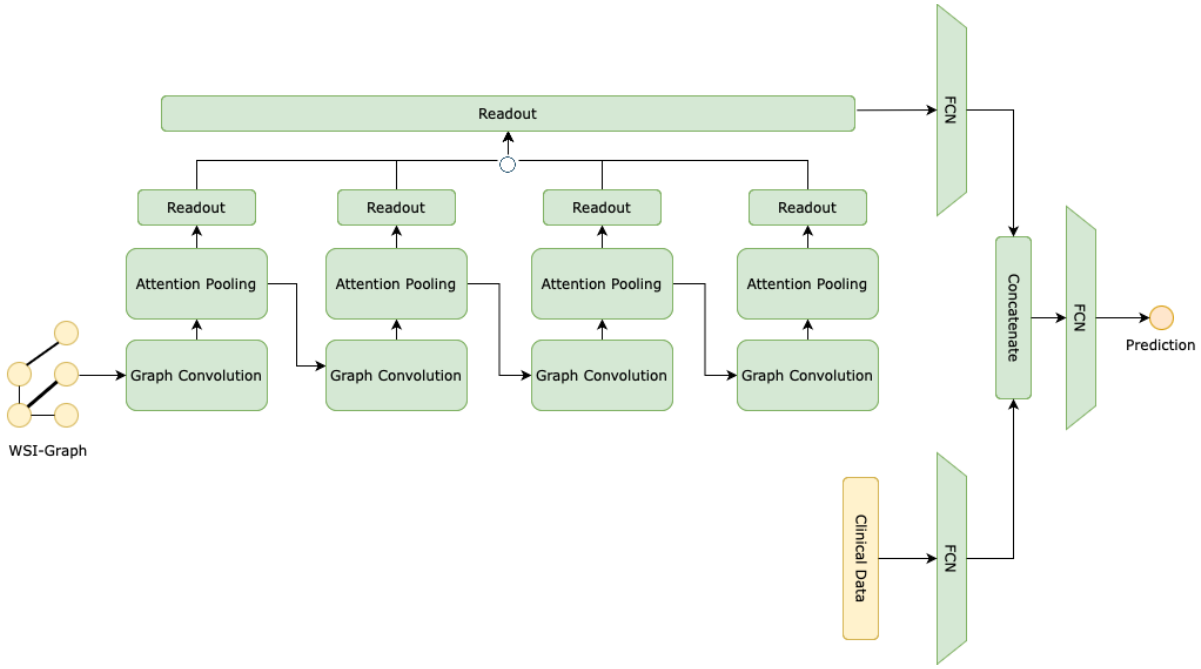


We used stochastic gradient descent (SGD) with learning rate of 1e-3 to train GNPC. A batch size of 64 and a maximum epoch of 100 were employed during training. Additionally, to prevent overfitting and improve generalisation, L2 regularisation with a coefficient of 1e-4, weight decay of 1e-5 and early stopping were applied.

#### *Model selection*

To ensure the generalisability and robustness of our model, we trained it on the SYSUCC cohort and evaluated its performance on unseen data from both the SYSUCC (internal test set) and CUHK (external test set) cohorts. We split the larger SYSUCC cohort into a discovery set (80%) and an internal testing set (20%), stratified by survival time across each survival endpoint. The entire CUHK cohort was reserved for external testing, providing an independent dataset to validate the model's generalisability.

To optimise model hyperparameters, we conducted 5-fold cross-validation on the discovery set. Models from the cross-validation study were evaluated using the C-index and the log-rank test of the Kaplan-Meier (KM) curve. For KM curve evaluation, we used the median risk score from the training set to stratify patients in the test set into low- and high-risk groups. We reported the mean and standard deviation of the C-index, as well as the median p-value from the log-rank test.

**Fig. S2 Model selection**

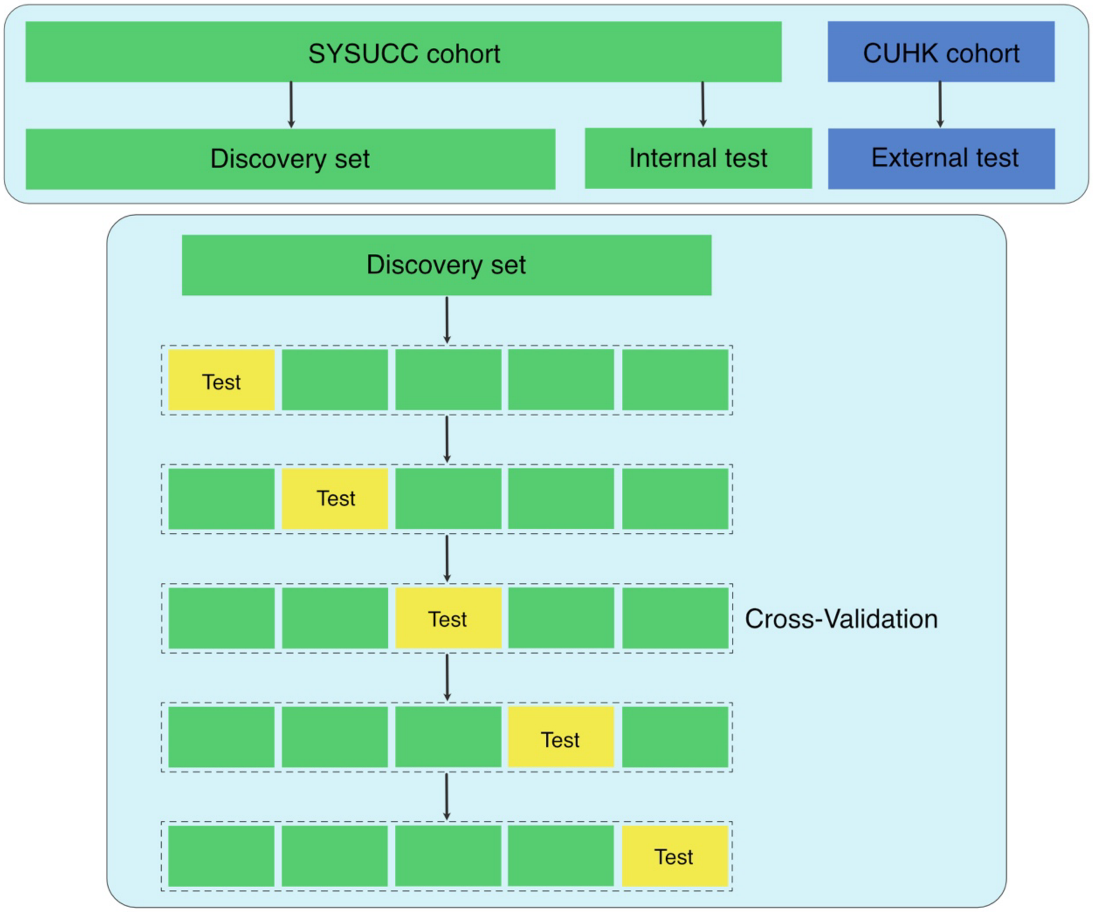


For final model evaluation, we trained on the full discovery set for each survival endpoint and then tested the model on both the internal and external test sets. In both the internal and external test sets, we applied the median risk score from the discovery set to define risk groups. A p-value of $<0.05$ was used as the threshold for statistical significance in all tests.

**Table S1 Clinicopathological characteristics of cohorts**

| Covariates | Sub-covariates | SYSUCC* | CUHK* |
|---|---|---|---|
| Age | | 46.10 (10.97) | 52.79 (12.47) |
| Sex | Female | 497 (27%) | 24 (24%) |
| | Male | 1352 (73%) | 76 (76%) |
| T | 1 | 214 (12%) | 24 (24%) |
| | 2 | 240 (13%) | 17 (17%) |
| | 3 | 1039 (56%) | 37 (37%) |
| | 4 | 356 (19%) | 22 (22%) |
| N | 0 | 186 (10%) | 18 (18%) |
| | 1 | 714 (39%) | 32 (32%) |
| | 2 | 594 (32%) | 33 (33%) |
| | 3 | 355 (19%) | 17 (17%) |
| All Stage | I | 50 (3%) | 10 (10%) |
| | II | 205 (11%) | 16 (16%) |
| | III | 942 (51%) | 37 (37%) |
| | IV | 652 (35%) | 37 (37%) |
| EBVDNA | ≤ 4,000 | 1044 (56%) | - |
| | > 4,000 | 805 (44%) | - |
| LMP1 | ≤ 100 | - | 72 (74%) |
| | > 100 | - | 25 (26%) |

* mean (std) for numerical data or number of count (percentage) for categorical data

#### Fig. S3 Consort diagram

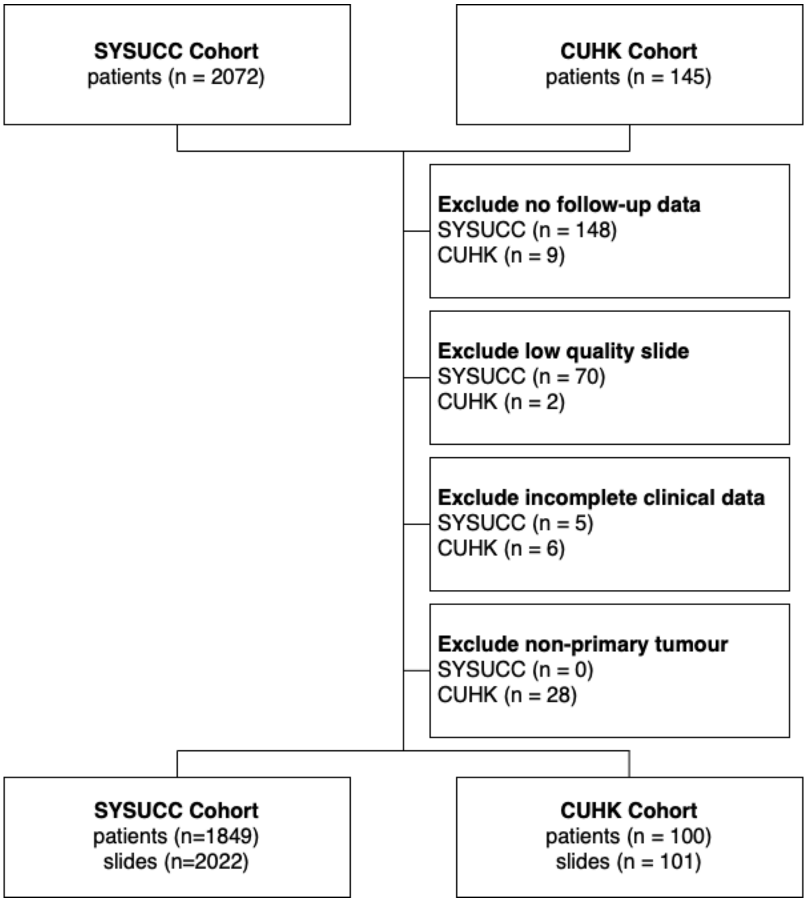

#### Fig. S4 KM-curves

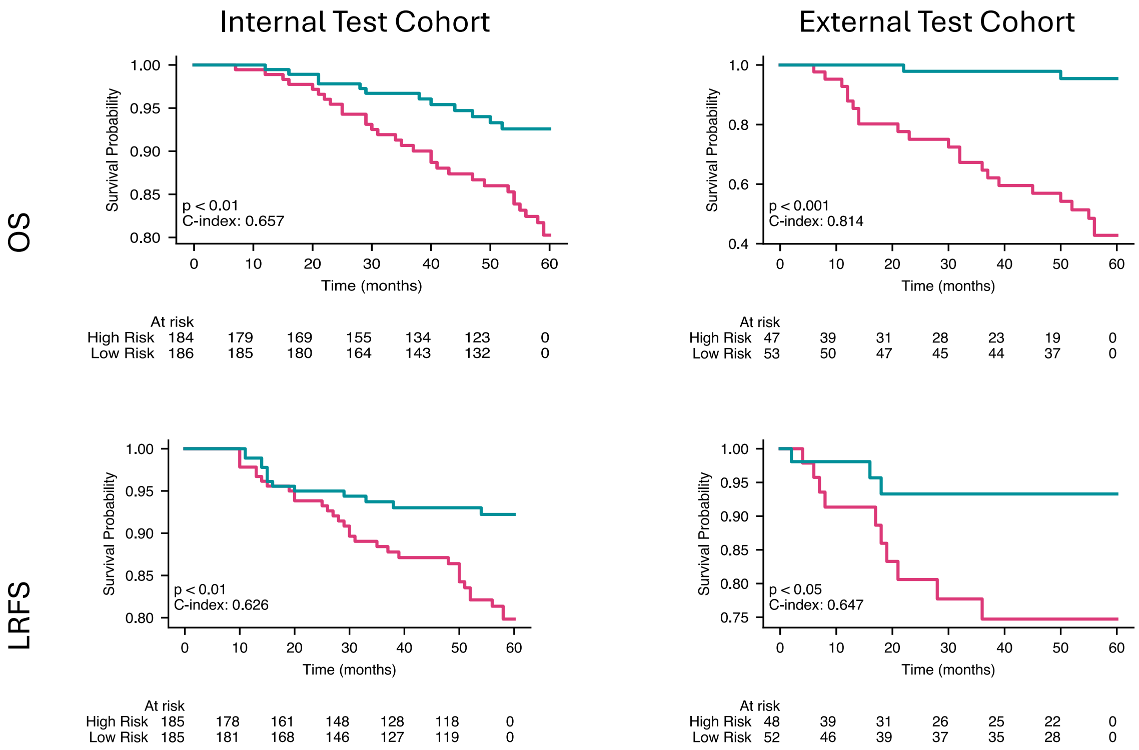

### Fig. S5 Cox-PH analysis on overall survival cases

#### Univariate Analysis

##### Internal Test Cohort

| Factor | Subgroup | HR (CI 95%) | p-value |
|---|---|---|---|
| **Age** | years | 1.01 (0.99-1.03) | 4.29e-01 |
| **Sex** | female | *reference* | |
| | male | 1.14 (0.66-1.97) | 6.36e-01 |
| **Smoking** | no | *reference* | |
| | yes | 1.34 (0.80-2.27) | 2.68e-01 |
| **Family** | no | *reference* | |
| | yes | 0.64 (0.24-1.70) | 3.67e-01 |
| **T** | 1 | *reference* | |
| | 2 | 0.50 (0.18-1.34) | 1.66e-01 |
| | 3 | 1.28 (0.70-2.34) | 4.17e-01 |
| | 4 | 1.74 (0.87-3.45) | 1.15e-01 |
| **N** | 0 | *reference* | |
| | 1 | 0.90 (0.48-1.69) | 7.47e-01 |
| | 2 | 1.29 (0.70-2.40) | 4.15e-01 |
| | 3 | 1.59 (0.80-3.15) | 1.87e-01 |
| **All Stage** | I | *reference* | |
| | II | 0.52 (0.17-1.57) | 2.49e-01 |
| | III | 0.92 (0.48-1.76) | 7.91e-01 |
| | IV | 1.88 (0.97-3.63) | 5.99e-02 |
| **EBVDNA** | ≤ 4,000 | *reference* | |
| | > 4,000 | 2.20 (1.31-3.67) | < 0.01** |
| **GNPC** | risk score | 1.61 (1.13-2.28) | < 0.01** |

0 1 2 3 4

##### External Test Cohort

| Factor | Subgroup | HR (CI 95%) | p-value |
|---|---|---|---|
| **Age** | years | 1.01 (0.99-1.03) | 4.29e-01 |
| **Sex** | female | *reference* | |
| | male | 1.14 (0.66-1.97) | 6.36e-01 |
| **Smoking** | no | *reference* | |
| | yes | 1.34 (0.80-2.27) | 2.68e-01 |
| **Family** | no | *reference* | |
| | yes | 0.64 (0.24-1.70) | 3.67e-01 |
| **T** | 1 | *reference* | |
| | 2 | 0.50 (0.18-1.34) | 1.66e-01 |
| | 3 | 1.28 (0.70-2.34) | 4.17e-01 |
| | 4 | 1.74 (0.87-3.45) | 1.15e-01 |
| **N** | 0 | *reference* | |
| | 1 | 0.90 (0.48-1.69) | 7.47e-01 |
| | 2 | 1.29 (0.70-2.40) | 4.15e-01 |
| | 3 | 1.59 (0.80-3.15) | 1.87e-01 |
| **All Stage** | I | *reference* | |
| | II | 0.52 (0.17-1.57) | 2.49e-01 |
| | III | 0.92 (0.48-1.76) | 7.91e-01 |
| | IV | 1.88 (0.97-3.63) | 5.99e-02 |
| **EBVDNA** | ≤ 4,000 | *reference* | |
| | > 4,000 | 2.20 (1.31-3.67) | < 0.01** |
| **GNPC** | risk score | 1.61 (1.13-2.28) | < 0.01** |

0 1 2 3 4

#### Multivariate Analysis

##### Internal Test Cohort

| Factor | Subgroup | HR (CI 95%) | p-value |
|---|---|---|---|
| **Age** | years | 1.01 (0.98-1.03) | 5.63e-01 |
| **Sex** | female | *reference* | |
| | male | 1.12 (0.62-2.03) | 7.02e-01 |
| **Smoking** | no | *reference* | |
| | yes | 1.25 (0.71-2.22) | 4.38e-01 |
| **Family** | no | *reference* | |
| | yes | 0.70 (0.26-1.90) | 4.79e-01 |
| **T** | 1 | *reference* | |
| | 2 | 0.55 (0.20-1.56) | 2.62e-01 |
| | 3 | 1.34 (0.70-2.59) | 3.77e-01 |
| | 4 | 1.07 (0.48-2.37) | 8.71e-01 |
| **N** | 0 | *reference* | |
| | 1 | 0.96 (0.50-1.85) | 9.07e-01 |
| | 2 | 1.26 (0.65-2.44) | 4.86e-01 |
| | 3 | 0.86 (0.38-1.91) | 7.05e-01 |
| **All Stage** | I | *reference* | |
| | II | 0.77 (0.22-2.63) | 6.71e-01 |
| | III | 0.81 (0.39-1.67) | 5.64e-01 |
| | IV | 1.62 (0.75-3.50) | 2.23e-01 |
| **EBVDNA** | ≤ 4,000 | *reference* | |
| | > 4,000 | 1.82 (1.06-3.13) | < 0.05* |
| **GNPC** | risk score | 1.11 (0.72-1.72) | 6.44e-01 |

0 1 2 3 4

##### External Test Cohort

| Factor | Subgroup | HR (CI 95%) | p-value |
|---|---|---|---|
| **Age** | years | 1.01 (0.98-1.03) | 5.63e-01 |
| **Sex** | female | *reference* | |
| | male | 1.12 (0.62-2.03) | 7.02e-01 |
| **Smoking** | no | *reference* | |
| | yes | 1.25 (0.71-2.22) | 4.38e-01 |
| **Family** | no | *reference* | |
| | yes | 0.70 (0.26-1.90) | 4.79e-01 |
| **T** | 1 | *reference* | |
| | 2 | 0.55 (0.20-1.56) | 2.62e-01 |
| | 3 | 1.34 (0.70-2.59) | 3.77e-01 |
| | 4 | 1.07 (0.48-2.37) | 8.71e-01 |
| **N** | 0 | *reference* | |
| | 1 | 0.96 (0.50-1.85) | 9.07e-01 |
| | 2 | 1.26 (0.65-2.44) | 4.86e-01 |
| | 3 | 0.86 (0.38-1.91) | 7.05e-01 |
| **All Stage** | I | *reference* | |
| | II | 0.77 (0.22-2.63) | 6.71e-01 |
| | III | 0.81 (0.39-1.67) | 5.64e-01 |
| | IV | 1.62 (0.75-3.50) | 2.23e-01 |
| **EBVDNA** | ≤ 4,000 | *reference* | |
| | > 4,000 | 1.82 (1.06-3.13) | < 0.05* |
| **GNPC** | risk score | 1.11 (0.72-1.72) | 6.44e-01 |

0 1 2 3 4

#### Fig. S6 Cox-PH analysis on local recurrence cases

##### Univariate Analysis

###### Internal Test Cohort

| Factor | Subgroup | HR (CI 95%) | p-value |
|---|---|---|---|
| **Age** | years | 1.02 (0.99-1.04) | 1.72e-01 |
| **Sex** | female | *reference* | |
| | male | 1.05 (0.59-1.85) | 8.73e-01 |
| **Smoking** | no | *reference* | |
| | yes | 1.20 (0.71-2.02) | 4.94e-01 |
| **Family** | no | *reference* | |
| | yes | 1.33 (0.53-3.34) | 5.44e-01 |
| **T** | 1 | *reference* | |
| | 2 | 0.58 (0.23-1.46) | 2.45e-01 |
| | 3 | 1.17 (0.65-2.09) | 6.00e-01 |
| | 4 | 1.29 (0.66-2.50) | 4.59e-01 |
| **N** | 0 | *reference* | |
| | 1 | 1.45 (0.80-2.61) | 2.16e-01 |
| | 2 | 1.00 (0.52-1.91) | 9.99e-01 |
| | 3 | 0.95 (0.46-1.96) | 8.90e-01 |
| **All Stage** | I | *reference* | |
| | II | 0.89 (0.36-2.19) | 7.96e-01 |
| | III | 1.14 (0.61-2.14) | 6.77e-01 |
| | IV | 1.15 (0.61-2.17) | 6.73e-01 |
| **EBVDNA** | ≤ 4,000 | *reference* | |
| | > 4,000 | 1.57 (0.96-2.57) | 7.34e-02 |
| **GNPC** | risk score | 1.79 (1.17-2.72) | < 0.01** |

###### External Test Cohort

| Factor | Subgroup | HR (CI 95%) | p-value |
|---|---|---|---|
| **Age** | years | 1.01 (0.97-1.05) | 6.52e-01 |
| **Sex** | female | *reference* | |
| | male | 1.01 (0.33-3.03) | 9.92e-01 |
| **Smoking** | no | *reference* | |
| | yes | 1.28 (0.50-3.27) | 6.01e-01 |
| **Family** | no | *reference* | |
| | yes | 0.55 (0.18-1.72) | 3.08e-01 |
| **T** | 1 | *reference* | |
| | 2 | 1.43 (0.38-5.35) | 5.93e-01 |
| | 3 | 0.72 (0.21-2.54) | 6.13e-01 |
| | 4 | 3.26 (1.03-10.25) | < 0.05* |
| **N** | 0 | *reference* | |
| | 1 | 1.48 (0.47-4.69) | 5.03e-01 |
| | 2 | 1.58 (0.50-4.97) | 4.38e-01 |
| | 3 | 1.10 (0.27-4.49) | 8.90e-01 |
| **All Stage** | I | *reference* | |
| | II | 1.22 (0.27-5.57) | 7.99e-01 |
| | III | 0.94 (0.26-3.34) | 9.19e-01 |
| | IV | 2.11 (0.65-6.89) | 2.16e-01 |
| **LMP1** | ≤ 100 | *reference* | |
| | > 100 | 1.64 (0.60-4.44) | 3.33e-01 |
| **GNPC** | risk score | 1.91 (0.84-4.36) | 1.25e-01 |

##### Multivariate Analysis

###### Internal Test Cohort

| Factor | Subgroup | HR (CI 95%) | p-value |
|---|---|---|---|
| **Age** | years | 1.01 (0.99-1.03) | 2.81e-01 |
| **Sex** | female | *reference* | |
| | male | 1.07 (0.59-1.94) | 8.29e-01 |
| **Smoking** | no | *reference* | |
| | yes | 1.05 (0.61-1.81) | 8.66e-01 |
| **Family** | no | *reference* | |
| | yes | 1.56 (0.60-4.05) | 3.62e-01 |
| **T** | 1 | *reference* | |
| | 2 | 0.63 (0.24-1.66) | 3.52e-01 |
| | 3 | 1.01 (0.51-1.99) | 9.73e-01 |
| | 4 | 1.05 (0.47-2.34) | 9.03e-01 |
| **N** | 0 | *reference* | |
| | 1 | 1.38 (0.75-2.54) | 3.04e-01 |
| | 2 | 0.98 (0.50-1.91) | 9.56e-01 |
| | 3 | 0.82 (0.36-1.87) | 6.44e-01 |
| **All Stage** | I | *reference* | |
| | II | 1.11 (0.41-3.05) | 8.38e-01 |
| | III | 1.02 (0.50-2.09) | 9.61e-01 |
| | IV | 0.98 (0.47-2.07) | 9.65e-01 |
| **EBVDNA** | ≤ 4,000 | *reference* | |
| | > 4,000 | 1.56 (0.93-2.60) | 8.91e-02 |
| **GNPC** | risk score | 1.67 (1.05-2.65) | < 0.05* |

###### External Test Cohort

| Factor | Subgroup | HR (CI 95%) | p-value |
|---|---|---|---|
| **Age** | years | 1.01 (0.97-1.05) | 7.37e-01 |
| **Sex** | female | *reference* | |
| | male | 0.83 (0.23-3.01) | 7.81e-01 |
| **Smoking** | no | *reference* | |
| | yes | 1.16 (0.40-3.31) | 7.87e-01 |
| **Family** | no | *reference* | |
| | yes | 0.77 (0.21-2.76) | 6.84e-01 |
| **T** | 1 | *reference* | |
| | 2 | 1.59 (0.39-6.49) | 5.18e-01 |
| | 3 | 0.64 (0.16-2.50) | 5.17e-01 |
| | 4 | 2.28 (0.53-9.84) | 2.70e-01 |
| **N** | 0 | *reference* | |
| | 1 | 1.39 (0.40-4.90) | 6.06e-01 |
| | 2 | 1.33 (0.39-4.54) | 6.44e-01 |
| | 3 | 0.91 (0.20-4.08) | 8.98e-01 |
| **All Stage** | I | *reference* | |
| | II | 1.00 (0.18-5.61) | 9.96e-01 |
| | III | 1.11 (0.26-4.65) | 8.91e-01 |
| | IV | 1.33 (0.32-5.56) | 6.97e-01 |
| **LMP1** | ≤ 100 | *reference* | |
| | > 100 | 1.93 (0.63-5.87) | 2.48e-01 |
| **GNPC** | risk score | 1.44 (0.54-3.87) | 4.64e-01 |

#### Fig. S7 Heatmap risks factor in overall survival cases

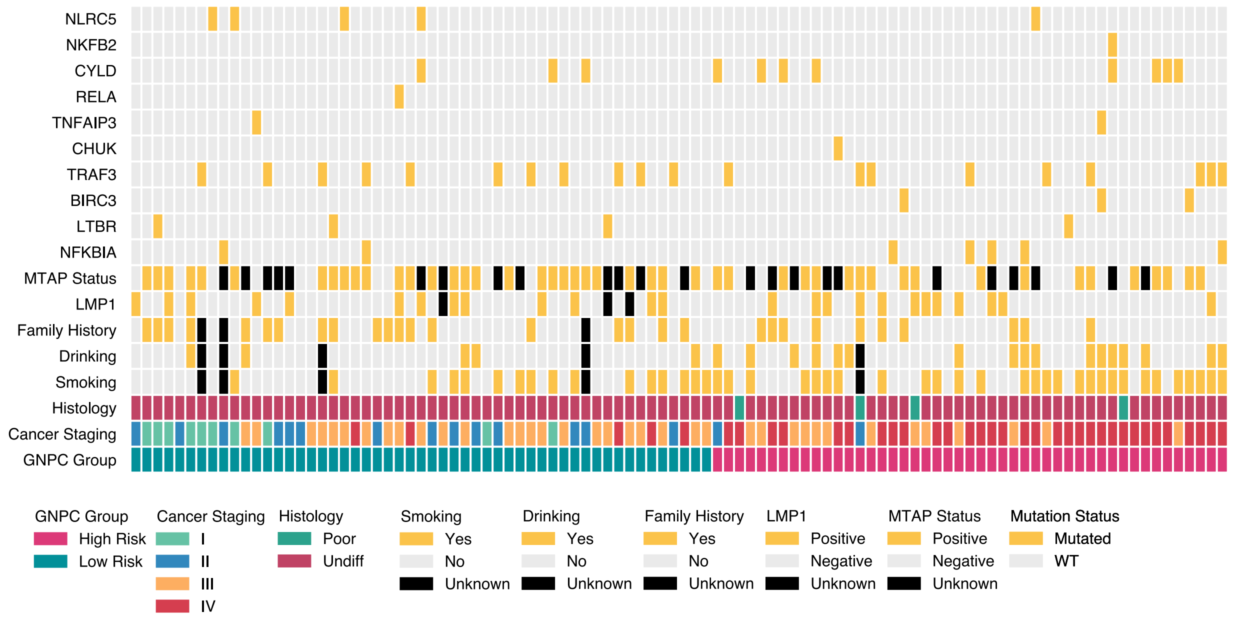


#### Fig. S8 Heatmap risks factor in local recurrence cases

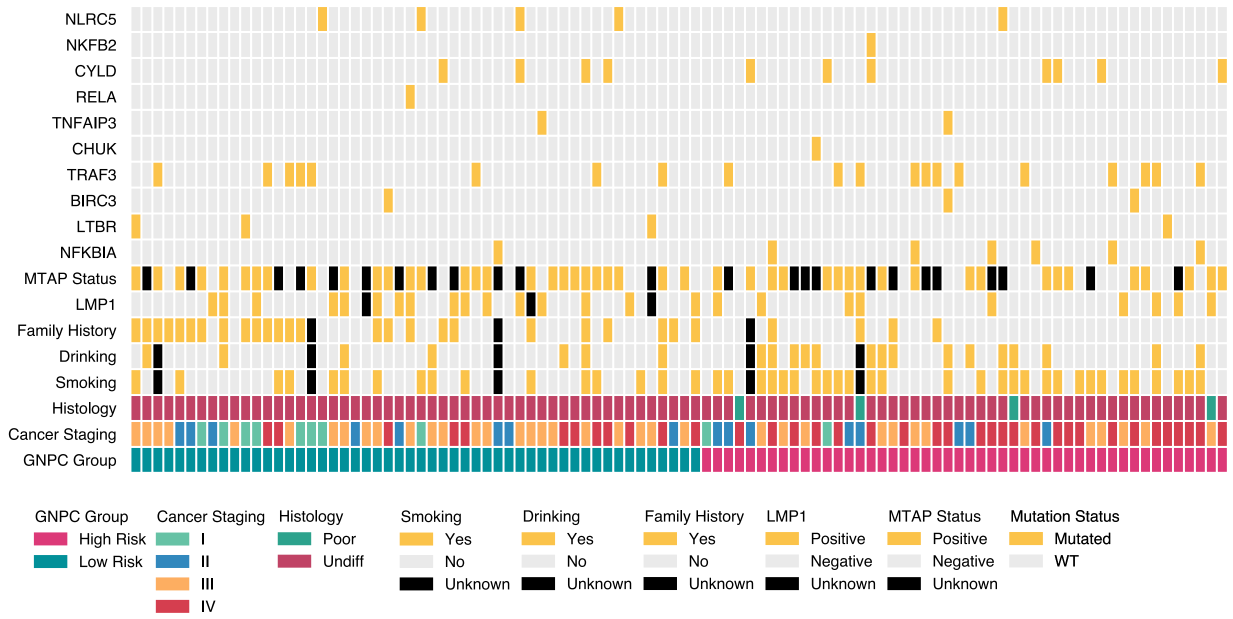

**Fig. S9 GNPC score association with LMP1 status**

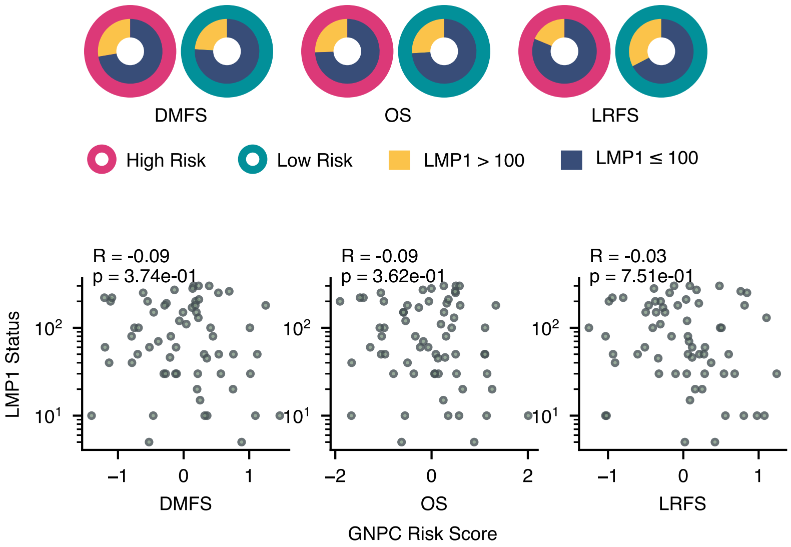
